# Determination of maximal oxygen uptake in adolescents

**DOI:** 10.1101/2025.08.27.25334536

**Authors:** Petri Jalanko, Emilia Laitinen, Dimitris Vlachopoulos, Ying Gao, Alan R. Barker, Bert Bond, Earric Lee, Eero A. Haapala

**Affiliations:** Sports & Exercise Medicine, Faculty of Sport and Health Sciences, University of Jyväskylä, Jyväskylä, Finland; Helsinki Clinic for Sports and Exercise Medicine, Foundation for Sports and Exercise Medicine, Helsinki, Finland; Children’s Health and Exercise Research Centre, Faculty of Health and Life Sciences, University of Exeter, Exeter, UK; Department of Sports Science, College of Education, Zhejiang University, Hangzhou, China; The Vascular Research Group, Faculty of Health and Life Sciences, University of Exeter; Montreal Heart Institute, Montréal, QC, Canada; School of Kinesiology and Physical Activity Sciences, University of Montréal, Montréal, QC, Canada; Institute of Biomedicine, University of Eastern Finland, Kuopio, Finland

## Abstract

**Purpose:** An oxygen uptake (V̇O_2_) plateau, despite an increased work rate, is considered the gold standard for confirming if exercise test performance reflects maximal oxygen uptake (V̇O_2max_). We investigated whether adolescents demonstrate a V̇O_2_ plateau during an incremental test or if a supramaximal verification phase is necessary to confirm V̇O_2max_. We also investigated the impact of using moving versus binned time averages on V̇O_2max_ values, and how these processing strategies influence the interpretation of the verification phase in confirming V̇O_2max_.

**Methods:** A total of 27 adolescents (16 girls) aged 12 to 14 years completed an incremental cycle ergometer ramp test to exhaustion. After a 15-minute recovery, a verification phase was conducted at 105% of their incremental test peak power. V̇O_2max_ was analysed using 15-second binned and moving averages.

**Results:** Out of 27 participants, 5 (19%) demonstrated a plateau in V̇O_2_ during an incremental test. V̇O_2max_ was confirmed in the verification phase for 23 out of the 27 adolescents (85%). The moving V̇O_2max_ (mL/kg/min) averages were higher than the binned V̇O_2_ values in the incremental test (1.8%) and the verification phase (2.4%) (P<0.0001). Processing strategies did not affect the confirmation of V̇O_2max_.

**Conclusion:** A verification phase is necessary for accurately determining V̇O_2max_ in adolescents, who often do not reach a V̇O_2_ plateau. The processing strategies of exercise tests should be reported, as different strategies can lead to variations in V̇O_2max_ results. However, these processing strategies do not impact the utility of the verification test.

## Introduction

Maximal oxygen uptake (V̇O_2max_) represents the upper physiological limit of the utilization of oxygen for producing energy during strenuous exercise performed until volitional exhaustion (Takken et al. 2017). V̇O_2max_ reflects the capacity of the respiratory, cardiovascular, and skeletal muscle systems to uptake, transport, and use oxygen and is considered the gold-standard measure of aerobic fitness (Armstrong and Welsman 1994). Accurate measurement of V̇O_2max_ is crucial as it is a valuable marker for determining cardiorespiratory fitness levels (Armstrong and Welsman 1994; Raghuveer et al. 2020), trainability of the cardiorespiratory system (McManus et al. 1997) and the development and severity of different health outcomes in children and adolescents (Pianosi et al. 2017). Methodological errors in measuring V̇O_2max_ can lead to inaccurate estimation and/or misclassification of aerobic fitness, which could affect health risk assessment and training prescriptions. However, uncertainty remains regarding the most accurate and valid method for measuring and interpreting V̇O_2max_ in children and adolescents.

In the laboratory, V̇O_2max_ is typically measured using a graded exercise test with an incremental ramp or step protocol until exhaustion (Myers et al. 1991). The presence of a V̇O_2_ plateau, despite an increased work rate, has been the most widely accepted method to assess whether the performance in the exercise test reflects the maximal effort and consequently, V̇O_2max_ (Niemeyer et al. 2021). However, it has been shown that only 20–60% of children and adolescents demonstrate a V̇O_2_ plateau during exercise tests (Rowland 1993; Armstrong et al. 1996; Barker et al. 2011; Niemeyer et al. 2021). In the absence of a V̇O_2_ plateau, the highest recorded V̇O_2_ is referred to as V̇O_2peak_. Secondary criteria are then used to verify maximal effort. However, these secondary criteria, such as heart rate and respiratory exchange ratio (RER), have been found to be insensitive and lack validity in determining whether the performance of the exercise test reflects the maximal effort of children and adolescents (Sansum et al. 2019). This led to the suggestion (Poole and Jones, 2017) that a subsequent supramaximal verification phase should be conducted to ascertain that V̇O_2max_ was indeed attained during the initial incremental test. V̇O_2max_ is considered confirmed if the increase in V̇O_2_ during the verification phase is less than 5% of the V̇O_2_ measured in the incremental test (Barker et al. 2011; Sansum et al. 2019). However, the evidence for using a verification phase in children and adolescents remains somewhat contentious and inconclusive.

Barker and colleagues (2011) examined healthy, recreationally active 9- to 10-year-old children and found that the verification phase confirmed the V̇O_2max_ measured during the incremental cycle ergometer ramp test in 12 out of 13 children. Lambrick and associates (2016) conducted a study among healthy, recreationally active children (9.6 ± 0.6 years old), using both discontinuous and continuous incremental treadmill tests to measure V̇O_2max_. Eight out of 21 participants achieved a higher V̇O_2max_ in the verification phase of the continuous incremental treadmill test compared to 4 out of 21 participants who achieved a higher V̇O_2max_ in the verification phase of the discontinuous incremental treadmill test. In contrast, Bhammar et al (2017) found that the verification phase did not confirm the V̇O_2max_ obtained from the incremental cycle ergometer test using a step protocol for any of the 23 children with and without obesity aged 10 to 12 years. The mean V̇O_2peak_ measured during the verification phase was 5–9% (0.10 to 0.14 L/min) higher than the mean V̇O_2peak_ obtained from the incremental cycle ergometer test (Bhammar et al. 2017). To the best of our knowledge, only one study has investigated whether a verification phase can confirm V̇O_2max_ in adolescents (Sansum et al. 2019). The authors found that the verification phase confirmed the V̇O_2max_ measured during the incremental cycle ergometer ramp test in 112 out of 128 children and adolescents aged 9 to 17 years. However, the results obtained by Sansum and colleagues (2019) have not been replicated in subsequent research involving adolescents in different laboratories.

The current clinical recommendations for pediatric exercise testing do not address the importance of V̇O_2_ processing strategies (Takken et al. 2017, 2025; Raghuveer et al. 2020). This oversight can significantly affect V̇O_2max_ results and clinical decision-making, potentially leading to participant misclassification and inappropriate treatment prescription. Additionally, it may lead to the failure to identify children in the population who are at an elevated risk for cardiovascular disease (Lang et al. 2024). The processing strategy of V̇O_2_ data, such as using binned or moving averages, has been found to cause 1–10% differences in V̇O_2 max_ among adolescents and adults (Blanchard et al. 2019; Martin-Rincon and Calbet 2020). While moving V̇O_2_ averages are recommended over binned V̇O_2_ averages (Robergs et al. 2010; Nolte et al. 2023; Hesse et al. 2025), most studies in children and adolescents (Rowland 1993; Bhammar et al. 2017; Sansum et al. 2019; Haapala et al. 2023) have used binned V̇O_2_ averages or have not explicitly reported the processing strategy. Additionally, the impact of V̇O_2_ processing strategies on the achieved V̇O_2max_ in the verification phase, and the difference between V̇O_2max_ values in the incremental test and verification phase, have yet to be explored (Costa et al. 2024). This difference could impact the interpretation of results from the verification phase.

Therefore, we examined whether adolescents aged 12–14 demonstrate a V̇O_2_ plateau during an incremental cycle ergometer test and if a verification phase would confirm the V̇O_2max_ measured in the incremental ramp test among this age group. We hypothesized that most adolescents would not demonstrate a V̇O_2_ plateau, but that the subsequent verification phase would confirm their V̇O_2max_. Secondly, we investigated whether processing the V̇O_2_ data from the incremental ramp test or verification phase using moving time averages, as opposed to binned time averages, would yield different V̇O_2max_ values. We hypothesized that using moving time averages would result in higher V̇O_2max_ levels compared to binned time averages. Lastly, we explored the impact of different V̇O_2_ processing strategies on the difference between the V̇O_2max_ values obtained during the incremental ramp test, and the verification phase. We hypothesized that these different processing strategies would not affect the difference in V̇O_2max_values between the incremental ramp test and the verification phase, nor would they influence the determination of whether V̇O_2max_ was confirmed.

## Methods

### Study population

The present analyses are based on the baseline assessment of the Vascular and Brain Health, Exercise, and Nutrition in Adolescents (VERNA) study (ISRCTN11803379). Altogether 28 adolescents (17 girls) aged 12–14 from the City of Jyväskylä, Finland, participated in the baseline examinations, which included the assessment of body size and composition and V̇O_2max_. The criteria for selecting the participants were: (1) aged 12–14 years and (2) apparently healthy. The health status was confirmed using a standardized physical activity readiness questionnaire. The exclusion criteria were: 1) Participation in another study that could affect the results 2) Use of supplements/medication affecting metabolism, vascular, and cognitive functions 3) History of metabolic or vascular disease, musculoskeletal injury or disability 4) Inability to understand study procedures 5) Relevant allergies or contraindications to exercise. The Research Ethics Committee of the Hospital District of Central Finland, Jyväskylä, Finland, approved the study protocol in 2022 (5U/2021). The parents or caregivers of the adolescents gave their written informed consent, and the adolescents gave their assent for participation. The VERNA study was carried out in accordance with the principles of the Declaration of Helsinki, which was revised in 2008. The funding sources had no role in the collection, analysis or interpretation of the data or publications.

### Incremental ramp test and verification phase protocol

We assessed V̇O_2_ during a maximal incremental ramp test using an electromagnetically braked Ergoselect 100 K^®^ cycle ergometer (Ergoline, Bitz, Germany) (Skog et al. 2020). The incremental ramp test protocol included a one-minute anticipatory period with the participant sitting on the ergometer; a 3-minute warm-up period with a workload of 20 watts; an exercise period with an increase in the workload of 1 watt per 3 seconds (20 W x min^-1^) until exhaustion, and a 2-minute recovery period with a workload of 20 watts. Participants were seated, and a rest period of 15 to 20 minutes followed before the commencement of the supramaximal verification phase. The verification phase began with a warmup of 3 minutes at 30 W. The resistance was then increased in a “step” transition to 105% of the peak power achieved in the incremental ramp test (Barker et al. 2011). The participants were required to cycle until exhaustion. Exhaustion was defined as the inability to maintain a cadence above 65 revolutions per minute in spite of vigorous verbal exhortation during both the incremental ramp test and verification phase. Following the verification phase, the participants completed a cooldown phase at 20 W. The participants were asked to keep the cadence stable and within 70–80 revolutions per minute. Heart rate was measured throughout the tests by a Polar H10 heart rate sensor (Polar Electro, Kempele, Finland) connected to the respiratory gas analyser.

### Assessment of oxygen uptake

V̇O_2_ (mL/min) was assessed by the Vyntus™ CPX Metabolic Cart (Vyaire Medical, USA). The respiratory gas analyser was calibrated for gas concentration and volume before each test according to the manufacturer’s recommendations. Respiratory gases were measured directly by the breath-by-breath method from the anticipatory period sitting on the ergometer to the end of the recovery period. The unmodified raw data was not filtered for outliers.

V̇O_2max_ from the incremental ramp test and the verification phase were defined using two different methods (Nolte et al. 2023). First, we recorded the highest binned 15-second V̇O_2_ value during the last minute of the test. We selected a 15-second average because this approach is more likely to accurately capture the true V̇O_2max_ without adding undue noise, given the short duration for which V̇O_2max_ can be sustained (Astorino 2009). Furthermore, previous studies involving children and adolescents have also employed the 15-second averaging method (Barker et al. 2011; Sansum et al. 2019). Subsequently, we manually searched the highest moving 15-second average V̇O_2_ value across the last minute of the test (SentrySuite V3.20.8, Vyaire Medical, USA). We normalised V̇O_2_ for body mass (BM) and skeletal muscle mass (SMM) to describe the differences between tests and data processing approaches using commonly used scaling approaches (Welsman and Armstrong 2019).

We classified the V̇O_2_ response as plateau (decelerated), linear, or accelerated using the V̇O_2_ – power output relationship during the linear portion of the test, excluding the first and last three minutes of the test (Day et al. 2003). The linear slope was then extrapolated and compared with residuals to determine the V̇O_2_ profile at exhaustion. Deviation from linearity during the last 3 min of exercise was determined by the difference between the extrapolated linear fit and the actual response at the point of fatigue. A plateau was defined as a negative residual ≥5% of the peak power projected V̇O_2_, an accelerated profile as a positive residual ≥5% of the projected V̇O_2_, and the linear response was defined as <5% of the peak power projected V̇O_2_ on either side of the extrapolated line (Sansum et al. 2019).

Using the same approach of prior studies (Barker et al. 2011; Sansum et al. 2019), we confirmed V̇O_2max_ when the increase in V̇O_2_ during the verification phase was less than 5% of the V̇O_2_ measured in the incremental ramp test, accounting for the typical within-participant error of measurement for V̇O_2max_ (Pivarnik et al. 1996).

### Assessment of body size, body composition, and somatic maturity

Body mass (BM), skeletal muscle mass (SMM), fat mass, and body fat percentage were measured with the adolescent having fasted for at least 3 hours, emptied the bladder, and standing in light underwear using a calibrated InBody® 770 bioelectrical impedance device (Biospace, Seoul, South Korea) to an accuracy of 0.1 kg. Body height was measured with the participant standing in the Frankfurt plane without shoes using a wall-mounted stadiometer to an accuracy of 0.1 cm. BMI (body mass index) was calculated by dividing weight (kg) by height (m) squared. ISO-BMI was computed based on Finnish reference values (Saari et al. 2011). ISO-BMI transforms age- and sex-specific child BMI to the corresponding adult BMI value. The prevalence of underweight (ISO-BMI/ BMI<17), normal weight (17–24.9), overweight (25–29.9), and obesity (≥30) was computed using standard cut-offs. Somatic maturity status in terms of time to peak height velocity was calculated using established equations (Moore et al., 2015).

### Statistical analyses

Statistical analyses were performed using the GraphPad Prism, version 9.5.1 (GraphPad Software, MA, USA). We used the medians and interquartile ranges or proportions to describe the characteristics of participants. We used paired samples t-tests for normally distributed variables to compare exercise responses measured during the incremental ramp test and the verification phase between binned and moving 15-second averages data processing strategies. We analysed the differences in the highest V̇O_2_ attained in the incremental ramp test (V̇O_2ramp_) and the verification phase (V̇O_2verification_) using binned and moving 15-second averages by the paired sample t-test. For skewed variables, we used the Wilcoxon matched pairs signed rank test with the method of Pratt for identical rows. A P value <0.05 was considered statistically significant. We also computed percentage differences for the abovementioned comparisons.

## Results

### Participant characteristics

28 participants attended the baseline visit, but one participant felt unwell after the incremental ramp test and did not perform the verification phase. Therefore, the final study sample included 27 adolescents (16 girls) aged 12–14 (Table 1). Because of device malfunction, valid heart rate recordings were only available for 19 adolescents. There were no differences in V̇O_2max_ normalised for BM or SMM between girls and boys (P>0.300) (Table 2). A total of 5 (18.5%), 18 (66.7%), and 4 (14.8%) participants demonstrated a plateau, linear, and accelerated V̇O_2_ profile, respectively from the initial incremental test (Table 2).

**Table 1.** Characteristics of participants.

| Background characteristics | All (n=27, females =16) |
| --- | --- |
| Age (years) | 12.9 (12.3 to 13.8) |
| Time to or from peak height velocity (years) | 0.4 (-0.2 to 1.2) |
| Body height (cm) | 159.7 (151.0 to 163.9) |
| Body mass (kg) | 50.6 (40.2 to 54.0) |
| BMI (kg / m<sup>2</sup>) | 19.0 (17.2 to 20.7) |
| ISO-BMI | 21.2 (19.9 to 23.0) |
| **Weight status (n/%)** |  |
| Underweight | 1 (3.7) |
| Normal weight | 23 (85.2) |
| Overweight | 3 (11.1) |
| Obese | 0 |
| Skeletal muscle mass (kg) | 20.4 (17.8 to 23.6) |
| Fat mass (kg) | 8.1 (6.5 to 12.9) |
| Body fat percentage (%) | 18.0 (14.3 to 23.9) |

Note. The data are median (interquartile range), or number of participants. BMI, Body Mass Index.

**Table 2.**
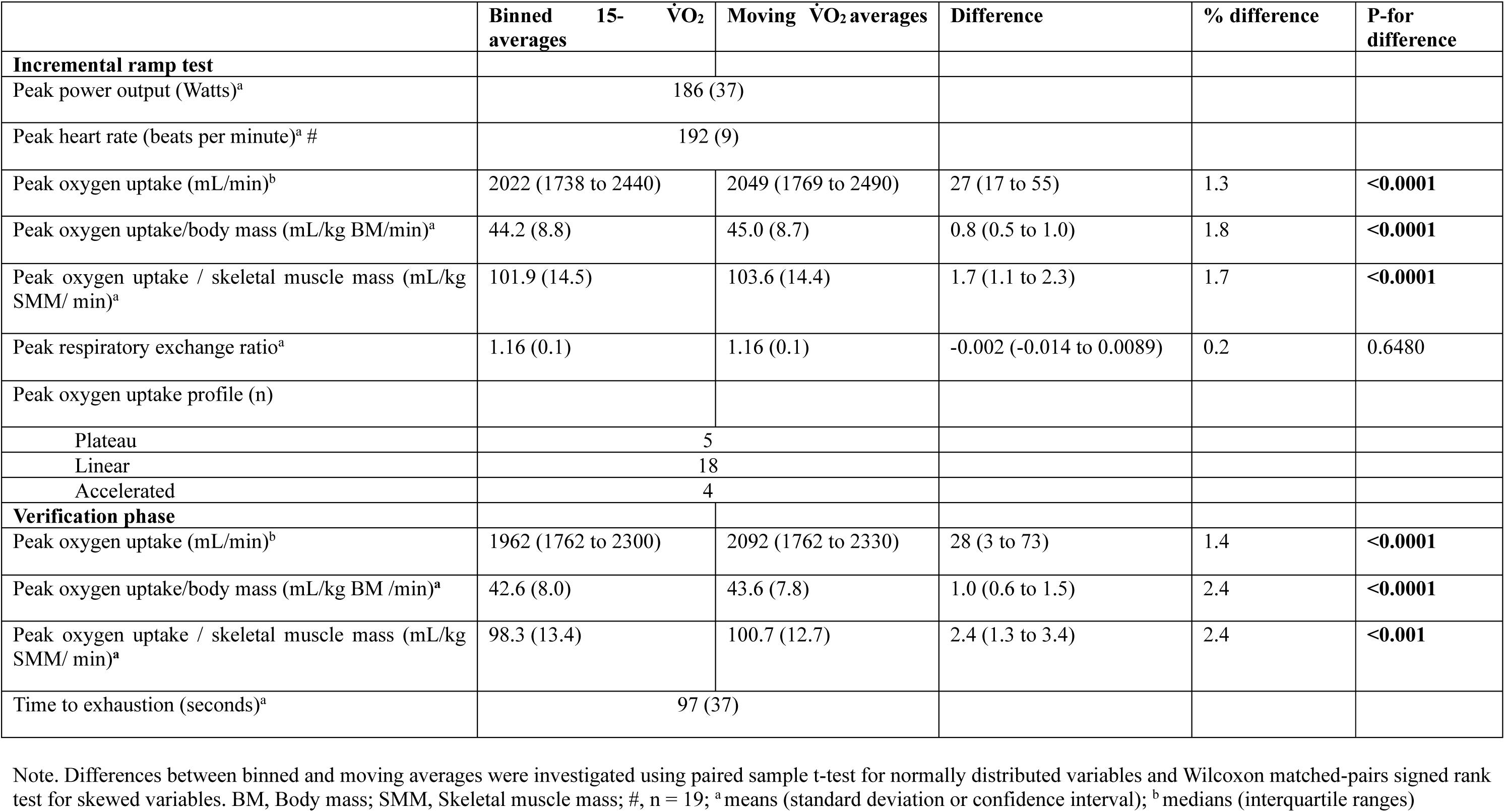
Characteristics of exercise test responses in 27 adolescents.

### Differences in oxygen uptake between the incremental ramp test and verification phase

Binned median of absolute V̇O_2verification_ (mean median difference -80 mL/min IQR=-199 to -3, p=0.0054, [3.3%, SD=6.6]), V̇O_2verification_ normalised for BM (mean difference -1.615 mL/kg BM/min, 95% CI=-2.690 to -0.5397, p=0.0048), and V̇O_2verification_ normalised for SMM (mean difference -3.581 mL/kg SMM/ min, 95% CI=-6.117 to -1.045, p=0.0075) were lower than corresponding values in the incremental ramp test (Figure 1 and 2; Table 2).

**Fig 1.**
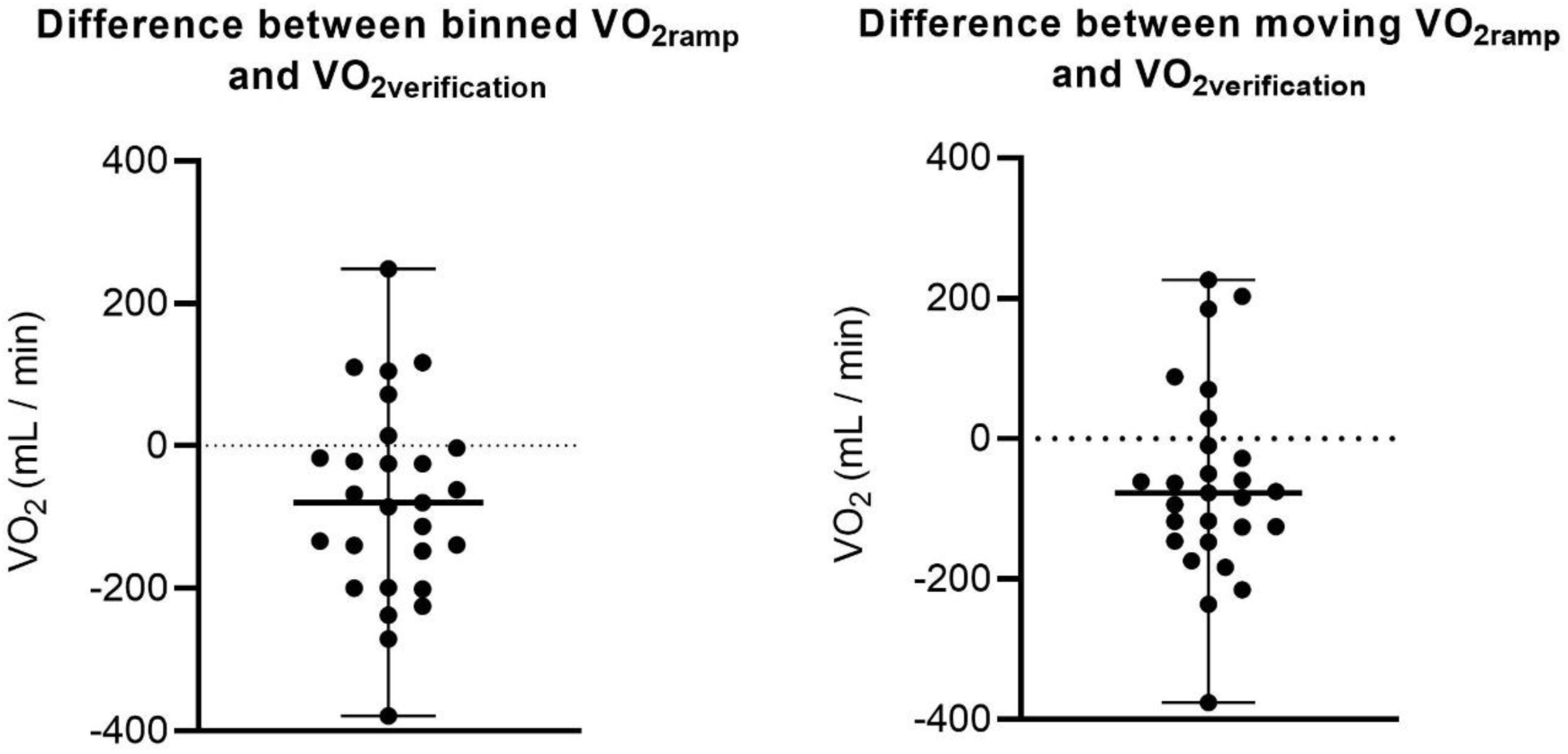
The difference between binned and moving averages in V̇O_2ramp_ and V̇O_2verification_.

**Fig 2.**
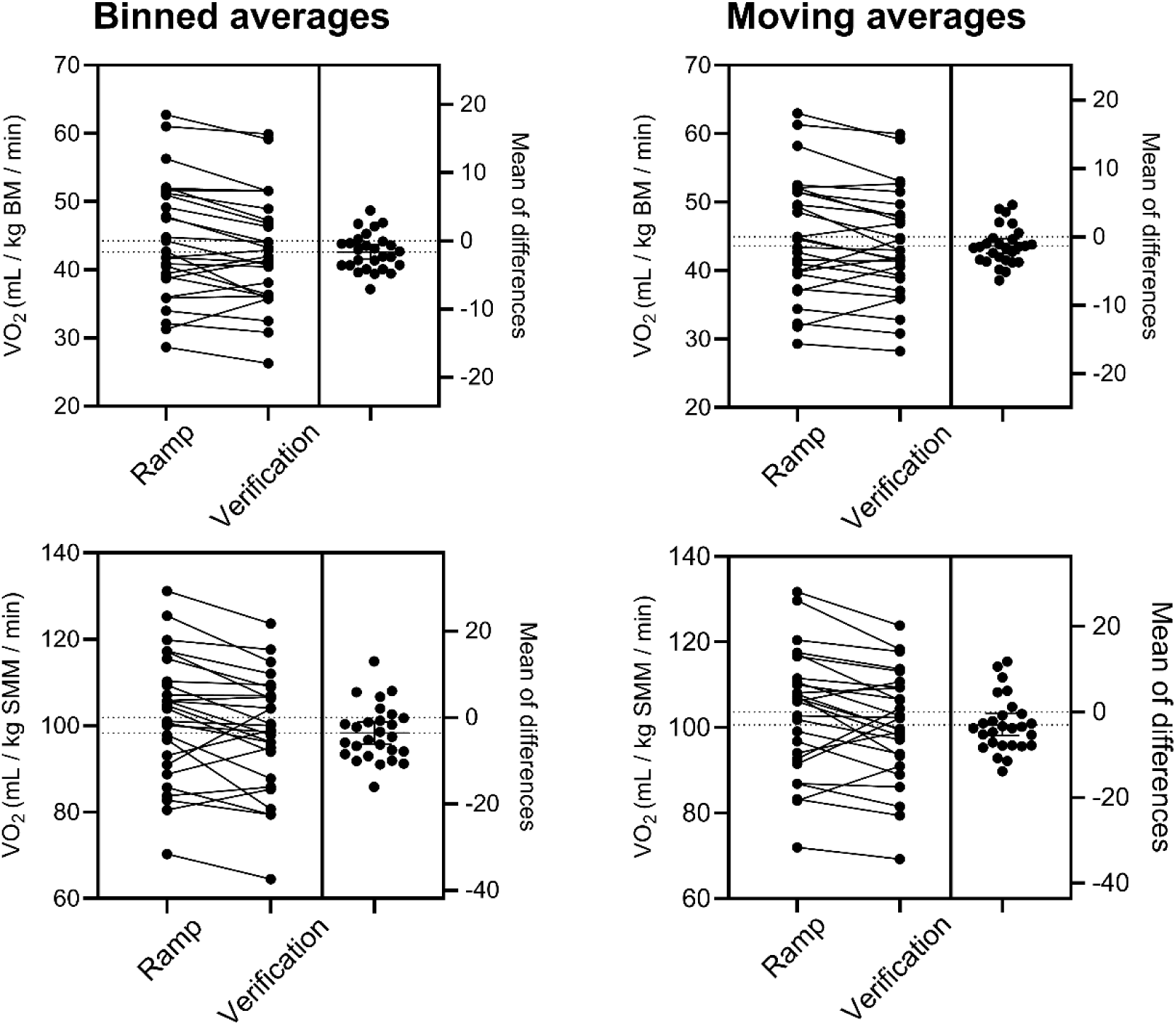
The difference between binned and moving averages in V̇O_2ramp_ and V̇O_2verification_ normalised for body mass and skeletal muscle mass. BM, Body mass; SMM, Skeletal muscle mass

Moving median of absolute V̇O_2verification_ (mean median difference -77 mL/min, IQR=-146 to -10, p=0.02, [2.5%, SD=6.4]), V̇O_2verification_ normalised for BM (mean difference -1.344 mL/kg BM/min, 95% CI=-2.441 to -0.2484, p=0.0182), and V̇O_2verification_ normalised for SMM (mean difference -2.996 mL/kg SMM/ min, 95% CI=-5.518 to -0.4139, p=0.0244) were lower than corresponding values in the incremental ramp test (Figure 1 and 2; Table 2).

Both the binned and moving averages of V̇O_2_ increased by more than 5% during the verification phase compared to the incremental ramp test for 4 out of 27 participants (15%), indicating that the verification phase confirmed V̇O_2max_ measured in the incremental ramp test for 23 out of 27 participants (85%).

### Differences between binned and moving oxygen uptake averages

In the incremental ramp test, the moving V̇O_2_ averages were higher than those of binned V̇O_2_ averages (Table 2). Similarly, in the verification phase, the moving V̇O_2_ averages were higher than those based on binned V̇O_2_ averages.

### Impact of the different processing strategies on the difference between the peak oxygen uptake values obtained during the incremental ramp test and the verification phase

A total of 6 participants had higher binned V̇O_2verification_ than V̇O_2ramp_ (Figure 1). From those six participants having higher binned averages in V̇O_2verification_ than V̇O_2ramp_, four (67%) participants were similarly classified by moving averages. From 21 participants having higher binned averages in V̇O_2ramp_ than V̇O_2verification_, 19 participants were similarly classified by moving averages.

Both binned and moving V̇O_2_ averages increased by >5% in the verification phase compared to the incremental ramp test in four (15%) participants, indicating that the verification phase did not confirm the V̇O_2ramp_ for those participants. Different V̇O_2_ processing strategies did not affect the differences between the V̇O_2max_ values obtained during the incremental ramp test and the verification phase. Therefore, the confirmation of V̇O_2ramp_ via the verification phase was unaffected by processing strategies.

### Characteristics between oxygen uptake profiles

One in five adolescents (20%) with a decelerated V̇O_2_ profile had higher moving V̇O_2_ in the verification phase than in the incremental ramp test. Four of 18 adolescents (22%) with linear profile had higher moving V̇O_2_ in a verification phase than in the incremental ramp test. One in four adolescents (25%) with an accelerated V̇O_2_ profile had higher moving V̇O_2_ in the verification phase than in the incremental ramp test. In four (15%) participants, V̇O_2_ increased >5% in the verification phase compared to the incremental ramp test, suggesting that the verification phase did not confirm the V̇O_2max_ measured during the incremental test. Of them, one had a plateau V̇O_2_ profile, two had linear V̇O_2_ profiles, and one had an accelerated V̇O_2_ profile.

## Discussion

The V̇O_2_ plateau, despite an increased work rate, has been the most commonly used phenomenon for assessing maximal effort and V̇O_2max_ during exercise tests. However, the occurrence of the V̇O_2_ plateau in adolescents has been questioned. Thus, performing a verification phase has been suggested (Barker et al. 2011; Niemeyer et al. 2021). We investigated whether adolescents demonstrate a V̇O_2_ plateau during an incremental cycle ergometer ramp test or if a verification phase is necessary to confirm the V̇O_2max_ obtained from the initial incremental test. We also examined if analysing V̇O_2_ responses with different processing strategies such as moving time averages, rather than binned time averages, affects V̇O_2max_ values obtained in the exercise tests. Out of 27 participants, only 5 (18.5%) showed a V̇O_2_ plateau during the incremental ramp test. Among the 19 out of 22 adolescents who did not exhibit a V̇O_2_ plateau in the incremental ramp test, V̇O_2max_ was confirmed in the verification phase. Notably, one adolescent who demonstrated a V̇O_2_ plateau in the incremental ramp test achieved a higher V̇O_2_ during the verification phase, indicating that the verification phase did not confirm the V̇O_2max_ measured during the incremental test for the participant. During both the incremental ramp test and the verification phase, the moving V̇O_2_ averages were higher than the binned V̇O_2_ values. Nevertheless, different V̇O_2_ processing strategies did not affect the interpretation of the verification phase in confirming the V̇O_2max_ measured during the incremental test.

A total of 5 (18.5%), 18 (66.7%), and 4 (14.8%) adolescents demonstrated a plateau, linear, and accelerated V̇O_2_ profile, respectively. This is somewhat similar to the findings of an earlier study (Sansum et al., 2019), which reported 35 (27%), 75 (59%), and 14 (18%) adolescents demonstrating a plateau, linear, and accelerated V̇O_2_ profile, correspondingly. The reason for the lack of a V̇O_2_ plateau, despite increasing exercise intensity, remains unclear. In their study, Sansum and colleagues (2019) investigated whether factors such as age, sex, or cardiorespiratory fitness could predict the absence of a V̇O_2_ plateau during incremental exercise tests in children and adolescents but found no significant predictors. Some evidence from studies on adults suggests that the absence of a V̇O_2_ plateau may be determined by physiological factors, such as low anaerobic capacity (Gordon et al. 2011), slow V̇O_2_ kinetics (Niemeyer et al. 2019), or excessive accumulation of metabolites during submaximal exercise (Lacour et al. 2007). The present study indicates that most adolescents do not display a V̇O_2_ plateau during the incremental ramp test. As such, whether performance in the incremental ramp test accurately represents maximal effort and V̇O_2max_ remains to be ratified. Taken together, these results suggest that it is essential to conduct a verification phase to confirm the V̇O_2max_ obtained from the incremental ramp test in adolescents.

The verification phase confirmed binned and moving V̇O_2max_ achieved in the incremental ramp test in 23 out of 27 participants, supporting our hypothesis. However, these findings contradict the results of an earlier study. Bhammar and coworkers (2017) investigated children aged 10 to 12 using an incremental cycle ergometer test and found that verification was unable to confirm V̇O_2max_. However, this could be due to methodological differences (step vs ramp protocol, Douglas bags vs breath-by-breath gas analyser). Furthermore, the study had an average time to exhaustion in the verification phase of 130 ± 63 s, which is longer than our average of 97 ± 37 s. This discrepancy may be attributed to an underestimation of the true peak power caused by an early termination of the incremental test in Bhammar and coworkers (2017) study. Such an underestimation can result in a longer time to exhaustion during the verification phase, which has been associated with a decreased likelihood of confirming the V̇O2max from the incremental test (Sansum et al. 2019).

Nevertheless, our results are in agreement with majority of the earlier studies in children (Barker et al. 2011; Lambrick et al. 2016) and adolescents (Sansum et al. 2019) with comparable methodologies. This suggests that the verification phase is able to ascertain the attainment of V̇O_2max_ during the incremental test for most participants. However, since the subsequent verification phase did not confirm the V̇O_2ramp_ for all participants, it may be necessary to conduct a secondary supramaximal verification phase either on the same day or on a separate day to confirm V̇O_2max_ in all adolescents adequately. It must be acknowledged, however, that this three-test protocol may not improve validity, or be feasible in all situations.

During both the incremental ramp test and the verification phase, the 15-second moving V̇O_2max_ averages were 0.8 ml/kg/min (1.8 %) and 1.0 ml/kg/min (2.4 %), respectively, higher than the 15-second binned V̇O_2_ values, supporting our hypothesis. Notably, there were significant interindividual variations, with one participant showing a moving V̇O_2ramp_ that was 2.8 ml/kg/min (6.8%) higher than the 15-second binned V̇O_2ramp_ (data not shown). Using binned time averages may lead to lower V̇O_2max_ values compared to moving averages, as the V̇O_2max_ may occur between two averaging intervals. This can result in an underestimation of V̇O_2max_ (Nolte et al. 2023). The 1 ml/kg/min difference between these processing strategies might be clinically significant for adolescents.

Hassinen and colleagues (2010) established that in adults, each 1 ml/kg/min increase in V̇O_2max_ is associated with a 0.19 unit decrease in z-scores for metabolic risk. Our findings support previous studies that reported 15-second moving V̇O_2max_ values to be between 0.9 and 1.7 ml/kg/min (1.7% to 3.2%) higher than the 15-second binned V̇O_2max_ values in well-trained adults during an incremental treadmill test (Scheadler et al. 2017). Similarly, in adolescents undergoing an incremental cycle ergometer test, the moving V̇O_2max_ values were found to be 0.7 ml/kg/min (1.7%) higher than the binned values. (Blanchard et al. 2019). We concur with Nolte et al. (2023) that data processing strategies which result in higher V̇O_2max_ values may not necessarily be more valid. However, our findings highlight the importance of reporting the specific V̇O_2max_ processing strategies used, as different strategies can lead to variations and incomparability in V̇O_2max_ results.

Future clinical recommendations for paediatric exercise testing should consider the significance of these processing strategies. Regardless, the V̇O_2_ processing strategies did not influence the interpretation of whether the verification phase confirmed the V̇O_2max_ measured during the incremental test. This suggests that researchers and practitioners can use either processing strategy when conducting incremental ramp test with a verification phase. Furthermore, in spite of the processing strategy employed, the conclusions drawn from the verification studies remain comparable.

There are some limitations to the present study that need to be considered. We calculated V̇O_2_ by manually averaging breath-by-breath data over 15-second intervals. Different approaches, such as averaging over 15 breaths, applying a digital filter, using varying time or breath averages, or employing a mixing chamber system, might yield different results (Robergs et al. 2010). Moreover, as we conducted an incremental ramp test on a cycle ergometer, the present findings might not apply to other protocols or exercise modalities. Nonetheless, the difference between binned and moving 15-second intervals has been demonstrated to be protocol-independent (Scheadler et al. 2017). Since we conducted only one verification phase at 105% of the peak power achieved during the incremental ramp test, we could not determine if the participants whose V̇O_2max_ was not confirmed would have had their V̇O_2max_ confirmed with different verification phase power levels or an additional verification phase. Lastly, participants were apparently healthy, of normal weight, and had moderate to high cardiorespiratory fitness. As a result, the findings cannot be generalized to clinical populations, individuals who are overweight, or those with low cardiorespiratory fitness levels.

In conclusion, only 5 out of 27 participants demonstrated a V̇O_2_ plateau during the incremental ramp test. However, the verification phase confirmed V̇O_2max_ in 19 of 22 adolescents who initially did not demonstrate a plateau in the incremental ramp test. This indicates that a verification phase could be utilized to accurately determine V̇O_2max_ in adolescents. Additionally, the moving V̇O_2max_ values were higher than the binned V̇O_2max_ values, indicating that different processing strategies may yield different V̇O_2max_ results. Therefore, processing strategies ought to be reported in detail for future studies. Future research is required to assess whether a different power level and the three-test protocol with a second verification phase would indeed be feasible to ascertain the V̇O_2max_ of participants, especially for those whose V̇O_2max_ remains unconfirmed during the initial verification phase. Furthermore, it is important to investigate whether varying the durations of sample time intervals, exercise modalities, or protocols would produce different results when comparing binned and moving time averages.

## Acknowledgements

The authors would like to thank all adolescents who participated in the VERNA study and the research group for their skilful contributions in performing these studies.

## Ethics approval statement

The Research Ethics Committee of the Hospital District of Central Finland, Jyväskylä, Finland, approved the study protocol in 2022 (5U/2021)

## Patient consent statement

Written informed consent was obtained from each adolescent’s parent or caregiver, and every adolescent provided assent to participation.

## Data availability

The datasets generated during and/or analyzed during the current study are not publicly available. The PI Dr. Eero Haapala can provide the data on reasonable request.

## Author Contribution Statement

P.J., E.L., D.V., A.R.B., B.B., EA.L., and E.AH. conceived and planned the experiments. E.L., EA.L., and E.A.H. carried out the experiments. P.J., E.L., D.V., Y.G., A.R.B., B.B., EA.L., and E.AH contributed to the interpretation of the results. E.A.H. designed the figures. P.J. took the lead in writing the manuscript. E.A.H. supervised the project. All authors provided critical feedback and helped shape the research, analysis and manuscript.

## Abbreviations

BM: Body mass
BMI: Body mass index
CRF: Cardiorespiratory fitness
RER: Respiratory exchange ratio
SMM: Skeletal muscle mass
V̇O_2_: Oxygen uptake
V̇O_2max_: Maximal oxygen uptake
V̇O_2peak_: Peak oxygen uptake

